# Gender and Racial/Ethnic Disparities in Pancreatic Cancer Incidence and Mortality in Texas: A Statewide Population-Based Analysis

**DOI:** 10.1101/2025.08.01.25332774

**Authors:** Ooreoluwa Fasola, Eunice Aregbesola, Gloria Erazo, Anjul Verma, Godfrey Tabowei, Foluke Atinuke Fasola

## Abstract

**Background:** Pancreatic cancer is the third leading cause of cancer-related deaths in the U.S. with rising incidence and mortality rates. Racial and ethnic disparities in pancreatic cancer outcomes in Texas have been underexplored. This study examined incidence and mortality of pancreatic cancer in Texas from 2000–2020.

**Methods:** We conducted a population-based analysis using data from the Texas Cancer Registry to assess incidence and mortality trends in pancreatic cancer by gender and race/ethnicity. Age-adjusted rates were calculated per 100,000 persons. Joinpoint Regression Program identified significant trends and calculated annual percentage change (APC).

**Results:** A total of 58,503 new cases and 48,692 deaths were identified during the study period. Males had higher incidence and mortality rates compared to females (13.9 vs. 10.8 and 11.9 vs. 9.0 per 100,000, respectively). Non-Hispanic Blacks had the highest incidence (16.4 per 100,000) and mortality (13.8 per 100,000) rates. Females demonstrated significantly increasing incidence rates (APC 1.21, p < 0.001) and mortality rates (APC 0.34, p < 0.001) during the study period. Incidence rates increased in Hispanics (APC 0.79, p < 0.001), non-Hispanics blacks (APC 0.45, p = 0.048) and fluctuating in non-Hispanic whites with a significant increase between 2009 -2018 (APC 2.17, p =0.011). Similarly, mortality rates increased in Hispanics (APC 0.78, p < 0.001), non-Hispanic whites (APC 0.63, p <0.05) and non-Hispanic Blacks (APC 0.46, p <0.05).

**Conclusions:** Increasing incidence and mortality trends, particularly among females and Hispanic populations, highlights the urgent need for targeted public health interventions, improved access to care, and early detection strategies.

## Introduction

Despite significant advancements in diagnostic and therapeutic medicine over the past few decades, pancreatic cancer remains one of the deadliest malignancies in the United States. It currently ranks as the third leading cause of cancer-related mortality, with projections estimating approximately 67 440 new cases in 2025 and a five-year relative survival rate of less than 15% [1]. Given the ongoing increase in both incidence and mortality rates, it is projected that pancreatic cancer will likely become the second leading cause of cancer deaths by 2030 [2]. Notably, emerging evidence indicates that these trends are not uniform and reveal substantial differences across gender and racial/ethnic groups [3].

Recent studies have shown that younger women, particularly those under 50 years of age across all major racial and ethnic categories, are experiencing disproportionately higher rates of pancreatic cancer incidence compared to their male counterparts with more pronounced increases observed among non-Hispanic Blacks [4]. Concurrently, non-Hispanic Black individuals develop and succumb to pancreatic cancer at significantly higher rates than those in other groups [5]. Additional research highlights clear disparities in mortality trends with mortality rates higher among non-Hispanic Blacks compared to other racial and ethnic groups [7].

However, these temporal trends in incidence and mortality across gender and ethnicity have been underexplored. Studying the population of Texas which is known for its large and ethnic diversity, this study analyzes temporal trends in both incidence and mortality across gender and ethnicity in Texas (2000-2020) using Texas cancer registry data and placing findings within context of broader national patterns. This aim was achieved. Understanding these disparities will help inform targeted public health interventions and future etiological research.

## Methods

The Texas Cancer Registry is a statewide and population-based cancer registry, and it is one of the largest cancer registries in the United States. It is Gold Certified by the North American Associations of Central Cancer Registries (NAACCR) [7]. We identified all patients diagnosed with pancreatic cancer as their primary malignancy from 2000 – 2020 from Texas Cancer Registry. Patients who died from pancreatic cancer were also identified from the TCR from 2000 – 2020. The ICD-0 codes used to identify pancreatic cancer cases include C250, C251, C252, C253, C254, C255, C256, C257, C258 and C259. The histology subtypes include 8000-8149, 8154, 8160-8231, 8243-8248, 8250-8682, 8690-8700, 8720-8790, 8971.

The primary outcome of interest was the incidence and mortality rates of pancreatic cancer from 2000 – 2020. The incidence rate is the number of new cancers of a specific site/type occurring in a specified population during a year, usually expressed as the number of cancers per 100,000 population at risk [7]. The mortality rate is the number of deaths, with cancer as the underlying cause of death, occurring in a specified population during a year. The independent variables of interest included in this study were gender and race/ethnicity. Gender was a binary variable with male and female. Race/ethnicity was categorized as non-Hispanic whites, non-Hispanic blacks, and Hispanics. Those outside these categories were excluded from the study.

### Statistical Analysis

We obtained data on annual incidence rates of pancreatic cancer cases per 100,000 persons. All ages (0 - 90+) were included in the study. The Texas Cancer Registry was accessed on Wednesday, March 2025. No authors had access to information that could identify participants during or after data collection. Texas Cancer Registry uses the direct standardization method to age-adjust pancreatic cancer incidence and mortality rates to the 2000 US standard population. The incidence and mortality rates were computed by gender and race/ethnicity. The trends in the incidence and mortality of pancreatic cancer by gender and race/ethnicity were analyzed by the Joinpoint Regression Program (Version 5.3.0.0) [8]. The Joinpoint Regression Program (Version 5.3.0.0) was produced by the Statistical Research and Applications Branch, National Cancer Institute. The program is useful in testing where a change in trend is statistically significant. The analysis starts with the minimum number of joinpoint and tests whether one or more joinpoints were statistically significant. We also used the Joinpoint regression software to calculate the annual percentage change (APC). Tests of significance use a Monte Carlo Permutation method [9]. The Monte Carlo permutation method helps test if there’s a real relationship between two variables by repeatedly permuting the data to break any real association, recalculating the statistic each time. The p-value is then computed as the proportion of permuted test statistics that are equal to or more extreme than the observed one, indicating how likely the observed result is under the null hypothesis.

### Ethics Statement

Institutional Review Board (IRB) approval was not required for this study. This research utilized data from the Texas Cancer Registry, which is a publicly available, population-based cancer surveillance database containing only de-identified patient information. The Texas Cancer Registry data are considered exempt from human subjects. research regulations under 45 CFR 46.104(d)(4) as the research involves the study of existing data that are publicly available and cannot be linked to specific individuals. No direct contact with patients or access to personally identifiable information occurred during this study. The research was conducted in accordance with the ethical principles outlined in the Declaration of Helsinki and followed all applicable guidelines for the use of population-based cancer registry data.

## Results

### Incidence

There was a total of 58,503 new cases of pancreatic cancer in Texas over a 21-year study period, i.e., 2000 - 2020. Table 1 shows the demographic characteristics of new cases of pancreatic cancer identified during the study period. Majority of pancreatic cancer cases were in males (n = 30,160). The highest incidence of pancreatic cancer was observed in the 70 -79 age group followed by the 60 - 69 age group. The overall age-adjusted incidence rates of pancreatic cancer were 12.2 per 100,000 persons, 13.9 per 100,000 males, 10.8 per 100,000 females, 11.6 per 100,000 Hispanics, 12.1 per 100,000 non-Hispanic whites, and 16.4 per 100,000 Non-Hispanic blacks respectively.

**Table 1.**
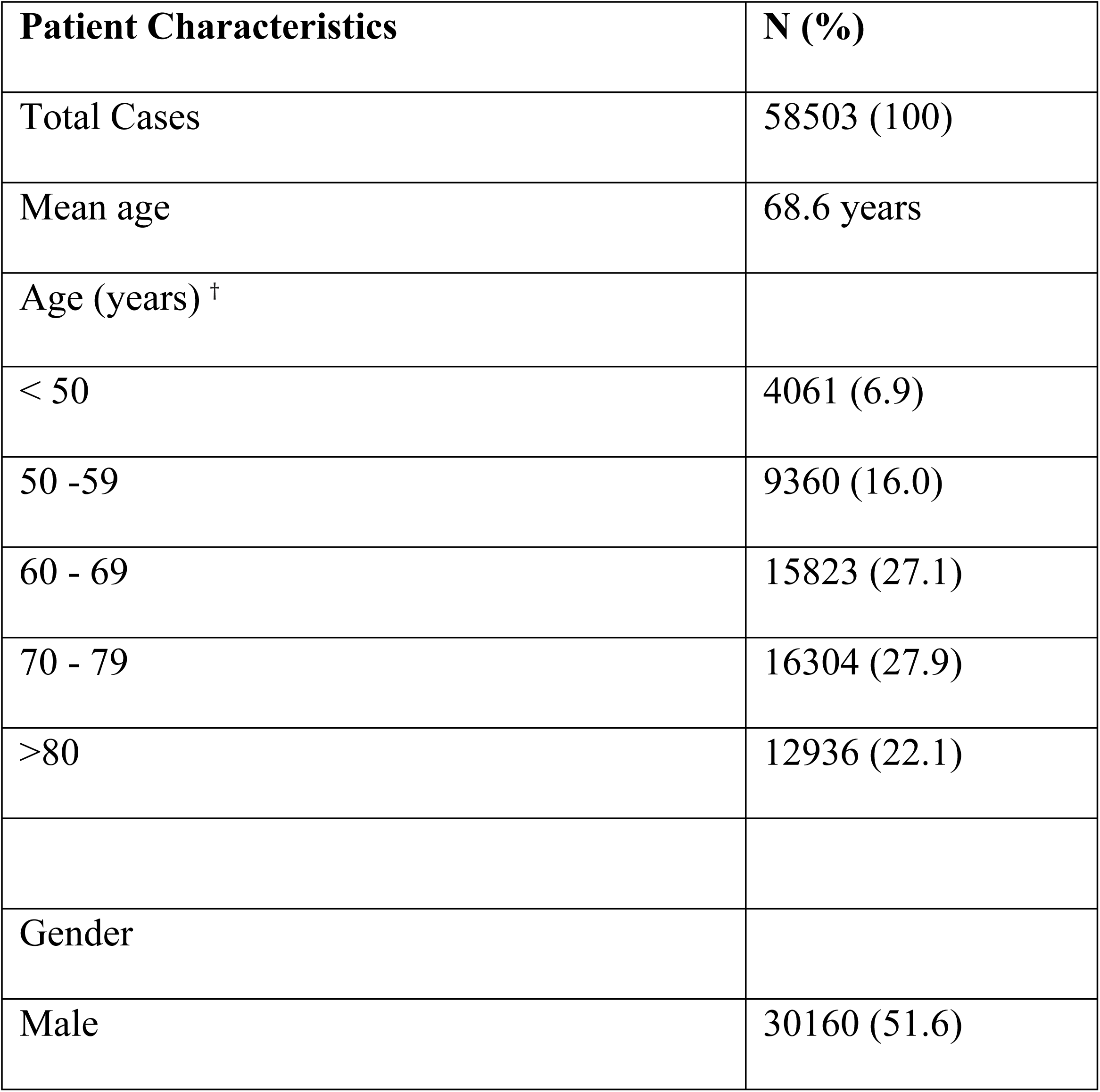
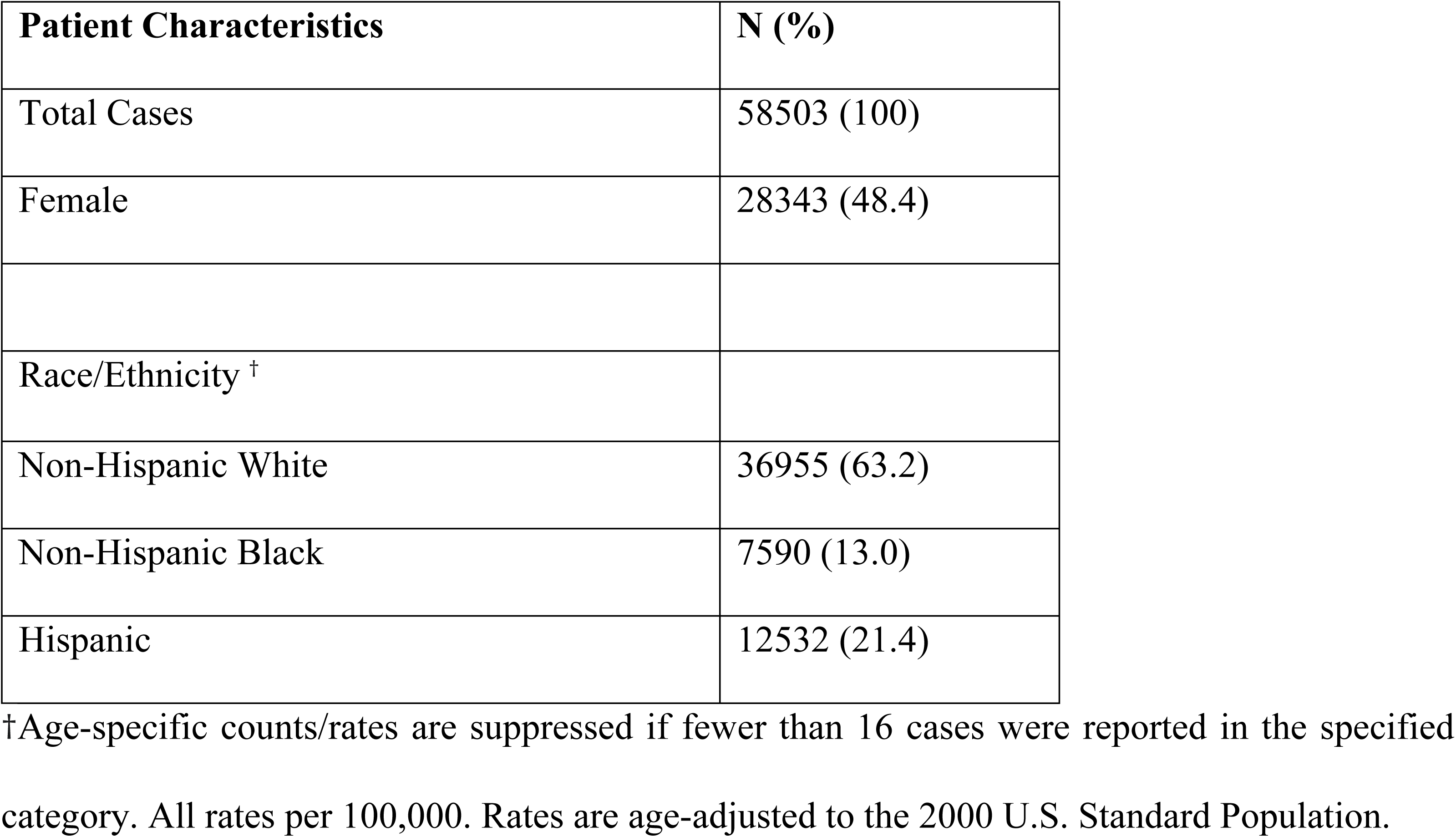
Demographic characteristics of new cases of pancreatic cancer in Texas between 2000 – 2020.

| Patient Characteristics | N (%) |
| --- | --- |
| Total Cases | 58503 (100) |
| Mean age | 68.6 years |
| Age (years) <sup>†</sup> |  |
| < 50 | 4061 (6.9) |
| 50 -59 | 9360 (16.0) |
| 60 - 69 | 15823 (27.1) |
| 70 - 79 | 16304 (27.9) |
| >80 | 12936 (22.1) |
| Gender |  |
| Male | 30160 (51.6) |
| Total Cases | 58503 (100) |
| Female | 28343 (48.4) |
| Race/Ethnicity <sup>†</sup> |  |
| Non-Hispanic White | 36955 (63.2) |
| Non-Hispanic Black | 7590 (13.0) |
| Hispanic | 12532 (21.4) |

<sup>†</sup>Age-specific counts/rates are suppressed if fewer than 16 cases were reported in the specified category. All rates per 100,000. Rates are age-adjusted to the 2000 U.S. Standard Population.

Fig 1 shows parallel comparisons of age-adjusted incidence trends from 2000 - 2020 by gender and race/ethnicity. Although, males had a higher overall incidence rate compared to female (13.9 vs 10.8 per 100,000 persons), the trends and changing patterns over the 10-year period were almost similar. Specifically, for males, pancreatic cancer incidence remained relatively stable from 2000 - 2003, then between 2003 - 2016, the rates fluctuated but generally on the rise. In 2016 however, the incidence was on the rise and decreased in 2020. Females experienced a similar fluctuating but steady rise in incidence rates until 2016 where the rates remained relatively stable.

**Fig 1.**
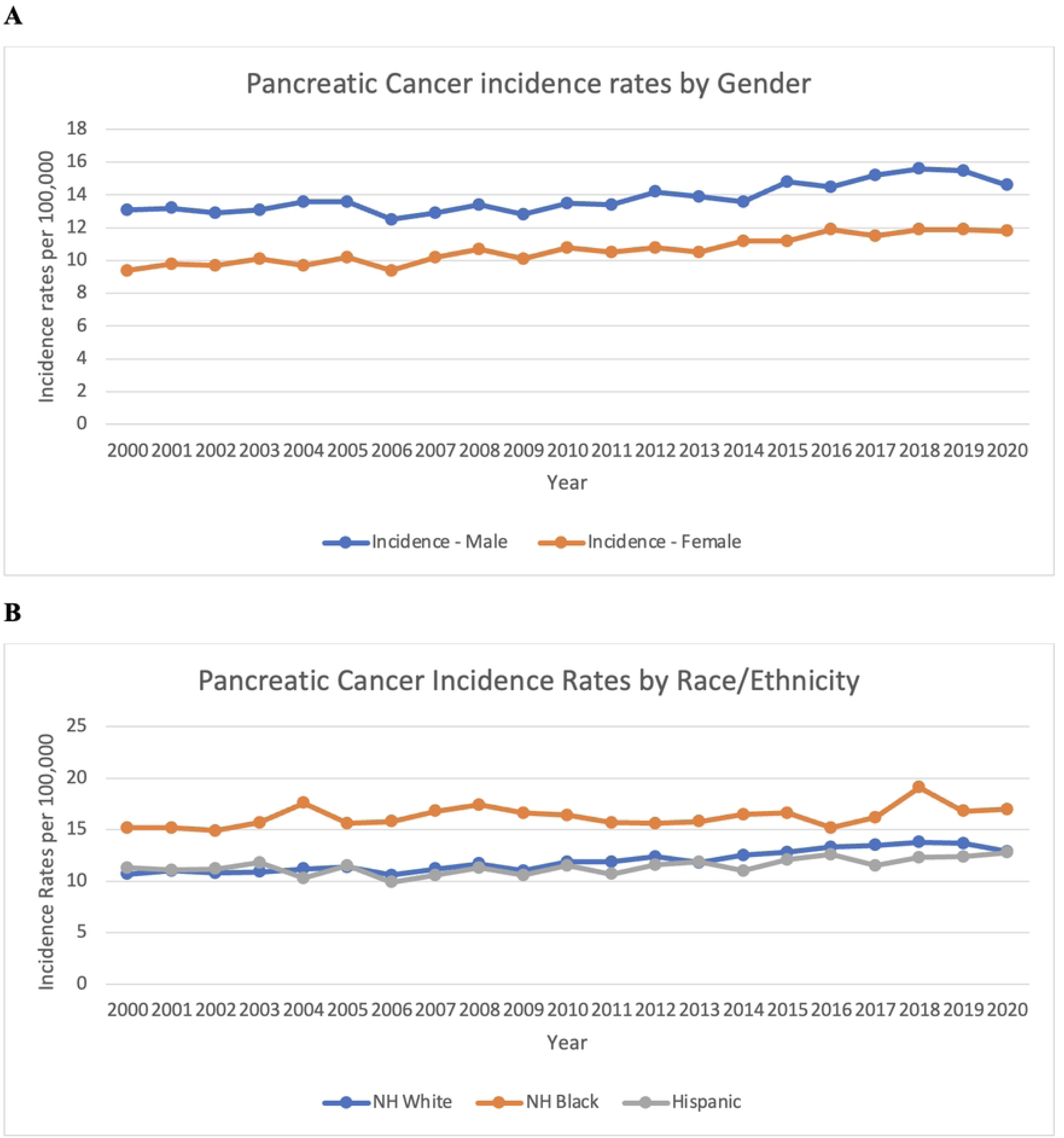
Trends in age-adjusted incidence rates of pancreatic cancer in Texas from 2000 – 2020. (A) Incidence rates by gender. (B) Incidence rates by race/ethnicity Joinpoint regression analysis of age-adjusted incidence rates of pancreatic cancer for males and females are shown in Fig 2. There was a statistically significant increasing trend between 2009 and 2020 (APC 1.47, 95% CI 0.40 - 4.71, p value < 0.05). For females however, there was a statistically significant increase in the incidence of pancreatic cancer from 2000 - 2020 (APC 1.21, 95% CI 0.98 - 1.48, p value < 0.001).

**Fig 2.**
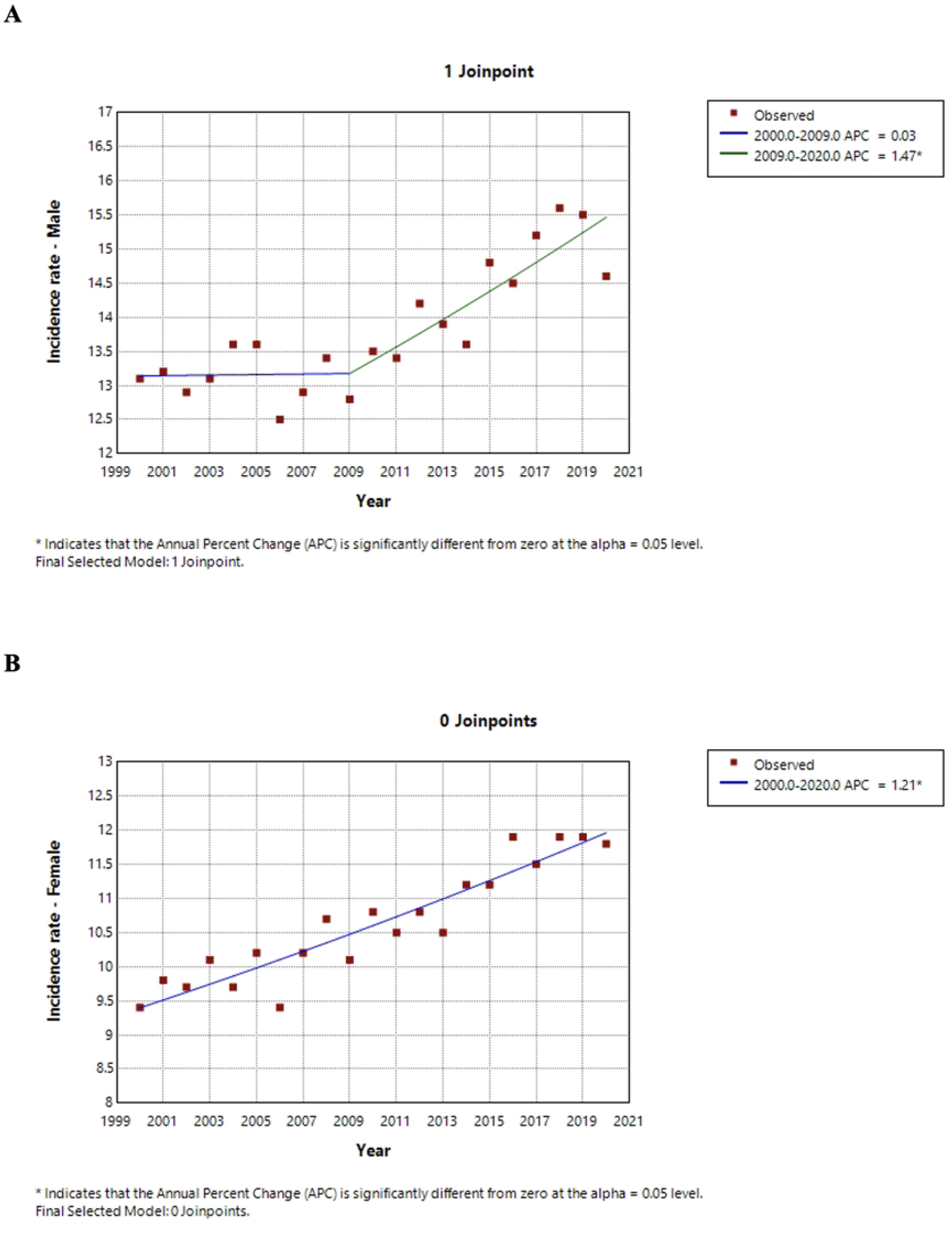
Joinpoint regression analysis of age-adjusted incidence rates of pancreatic cancer in Texas from 2000 -2020 by gender. (A) Age-adjusted incidence rates for males. (B) Age adjusted incidence rates for females. Joinpoint regression analysis of age-adjusted incidence rates of pancreatic cancer for Hispanics, non-Hispanic Blacks and non-Hispanic Whites are shown in Fig 3. The joinpoint regression analysis in terms of race/ethnicity revealed an increasing trend from 2000 - 2020 in the Hispanic patients which was statistically significant (APC 0.79, 95% CI 0.40 - 1.25, p value < 0.001 as well as Non-Hispanic Blacks (APC 0.45 95% CI 0.01 - 0.97, p value < 0.05). Interestingly, in Non-Hispanic whites, there was an increasing trend between 2000 - 2009 which was not significant. However, in 2009 - 2018, there was a statistically significant increase between 2009 - 2018 (APC 2.17, 95% CI 1.70 - 4.49, p value < 0.01) and a downward trend from 2018 - 2020 which was not statistically significant.

**Fig 3.**
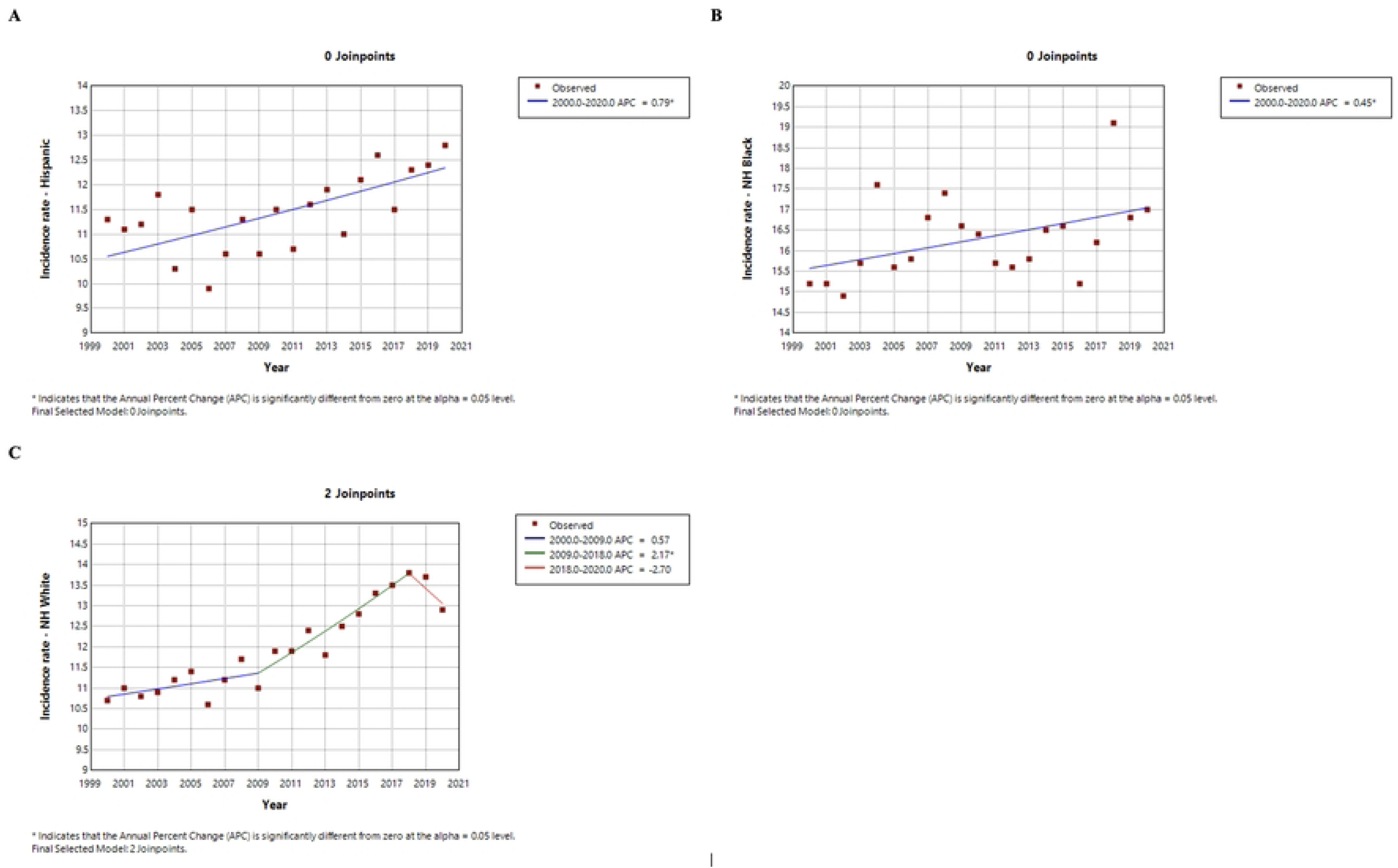
Joinpoint regression analysis of age-adjusted incidence rates of pancreatic cancer in Texas from 2000 -2020 by race/ethnicity. (A) Age-adjusted incidence rates for Hispanic (B) Age adjusted incidence rates for non-Hispanic Black. (C) Age-adjusted incidence rate for non-Hispanic Whites

Overall, Non-Hispanic Blacks had a higher incidence rate of pancreatic cancer over the study period. In Non-Hispanic Blacks, there was a steep increase between 2003 and 2004, and between 2017 and 2019. For Hispanics, the rates were stable early on, then fluctuated until 2017 where there was a decrease, afterwards remaining relatively stable. Non-Hispanic Whites had a stable but gradual increase in incidence rates over the period until 2019 here the rates decreased.

### Mortality

There were 48,692 cases of pancreatic cancer mortality in the same study period i.e., 2000 - 2020. Table 2 shows the demographic characteristics of deaths from pancreatic cancer identified during the study period. Mortality from pancreatic cancer was most prevalent among older adults, with the highest number of deaths occurring in patients aged 70–79, followed closely by those aged over 80. There was a slight male predominance in mortality. Racial and ethnic distribution revealed that Non-Hispanic White patients experienced the highest mortality, followed by Hispanic and Non-Hispanic Black patients.

**Table 2.** Demographic characteristics of pancreatic cancer decedents in Texas between 2000-2020.

| Patient Characteristics | N (%) |
| --- | --- |
| Total Cases | 48692 (100) |
| Mean age |  |
| Age (years) <sup>‡</sup> |  |
| < 50 | 2234 (4.6) |
| 50 -59 | 6774 (13.9) |
| 60 - 69 | 12594 (25.9) |
| 70 - 79 | 14261 (29.3) |
| > 80 | 12806 (26.3) |
| Gender |  |
| Male | 25127 (51.6) |
| Female | 23565 (48.4) |
| Race/Ethnicity ‡ |  |
| Non-Hispanic White | 31903 (65.5) |
| Non-Hispanic Black | 6202 (12.7) |
| Hispanic | 9398 (19.3) |

‡Age-specific counts/rates are suppressed if fewer than 16 cases were reported in the specified category. All rates per 100,000. Rates are age-adjusted to the 2000 U.S. Standard Population.

Fig 4 shows the trends in age-adjusted mortality rates by gender and race/ethnicity over the study period. Overall, the age-adjusted mortality rates in males were higher compared to females (11.9 vs 9.0 per 100,000 persons). In both males and females, the mortality rates remained relatively stable with a slight increase over time.

**Fig 4.**
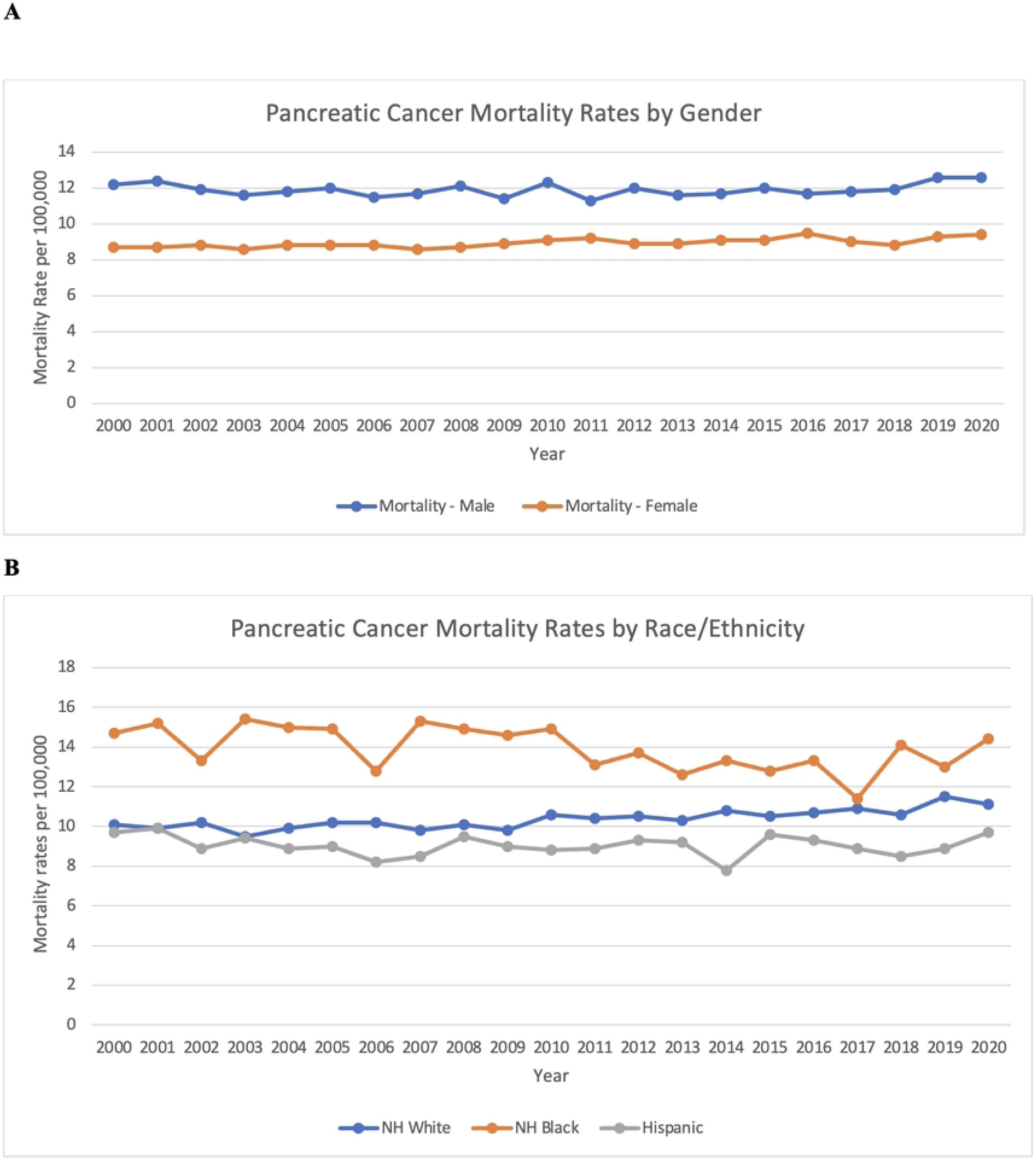
Trends in age-adjusted mortality rates of pancreatic cancer in Texas from 2000 – 2020. (A) Mortality rates by gender. (B) Mortality rates by race/ethnicity Joinpoint regression analysis of age-adjusted incidence rates of pancreatic cancer for males and females are shown in Fig 5. The analysis shows that there was a downward trend in the mortality rate of males from pancreatic cancer between 2000 - 2017 which was not statistically significant. However, from 2017 to 2020, there was a significant increase in the mortality rates (APC 2.76 95% CI 0.07 - 5.94, p value < 0.05). For females, there was a consistent upward trend in the mortality rates (APC 0.34 95% CI 0.18 - 0.51, p value < 0.01)

**Fig 5:**
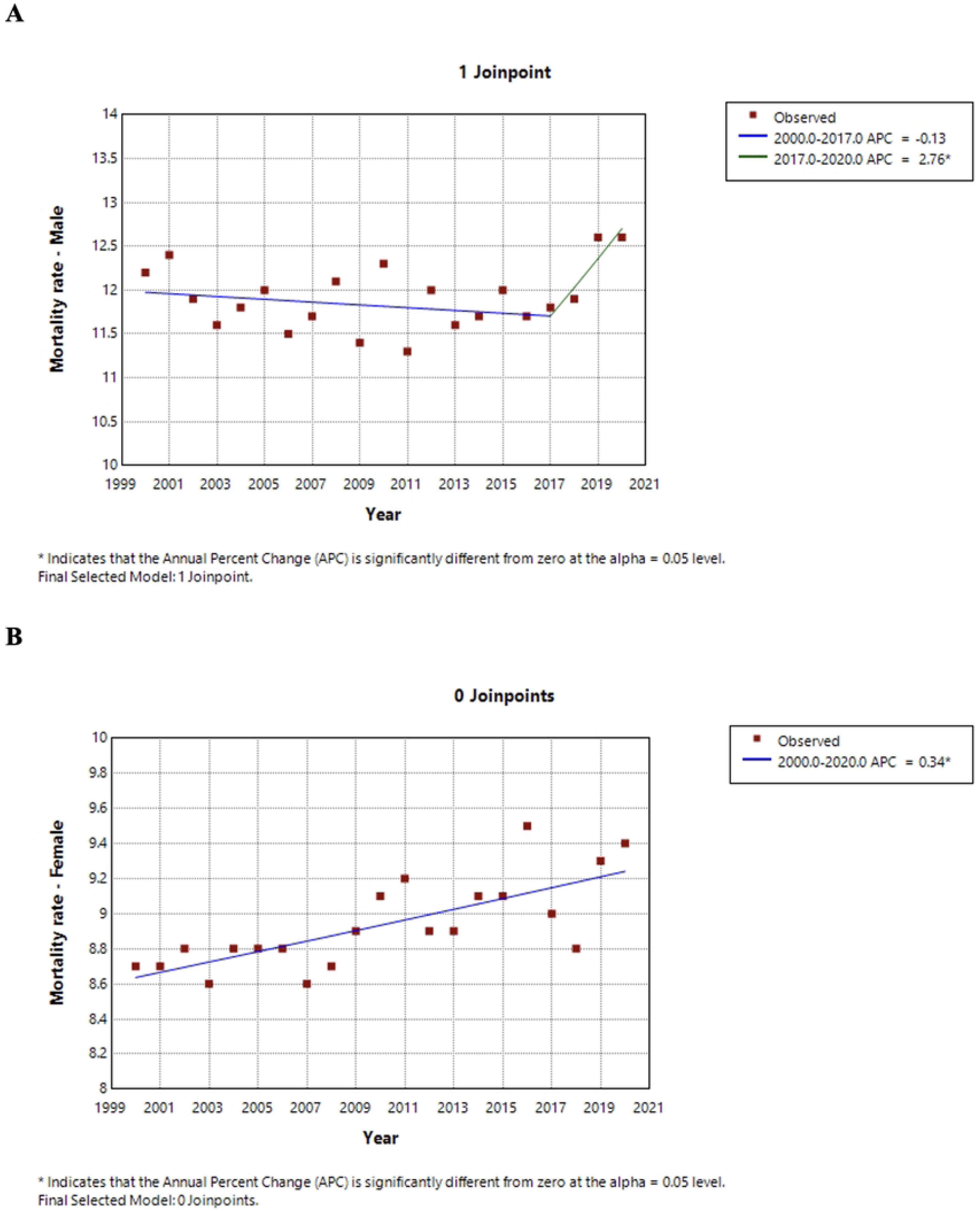
Joinpoint regression analysis of age-adjusted mortality rates of pancreatic cancer in Texas from 2000 -2020 by gender. (A) Age-adjusted mortality rates for males. (B) Age adjusted mortality rates for females. Joinpoint regression analysis of age-adjusted mortality rates of pancreatic cancer for Hispanics, non-Hispanic Blacks and non-Hispanic Whites are shown in Fig 6. The joinpoint regression analysis of the mortality rates in terms of race/ethnicity revealed an increasing trend from 2000–2020 in Hispanic patients, which was statistically significant (APC 0.78, 95% CI 0.40–1.25, p < 0.001), as well as in Non-Hispanic Blacks (APC 0.46, 95% CI 0.01–0.97, p < 0.05). The scatter of observed data points around the trend line is wider for Non-Hispanic Black individuals, suggesting greater annual variability compared to the other groups. Among Non-Hispanic Whites, there was a statistically significant increasing trend over the entire period from 2000 to 2020 (APC 0.63, p < 0.05), with a consistent upward trajectory in mortality rates without any significant changes in slope.

**Fig 6:**
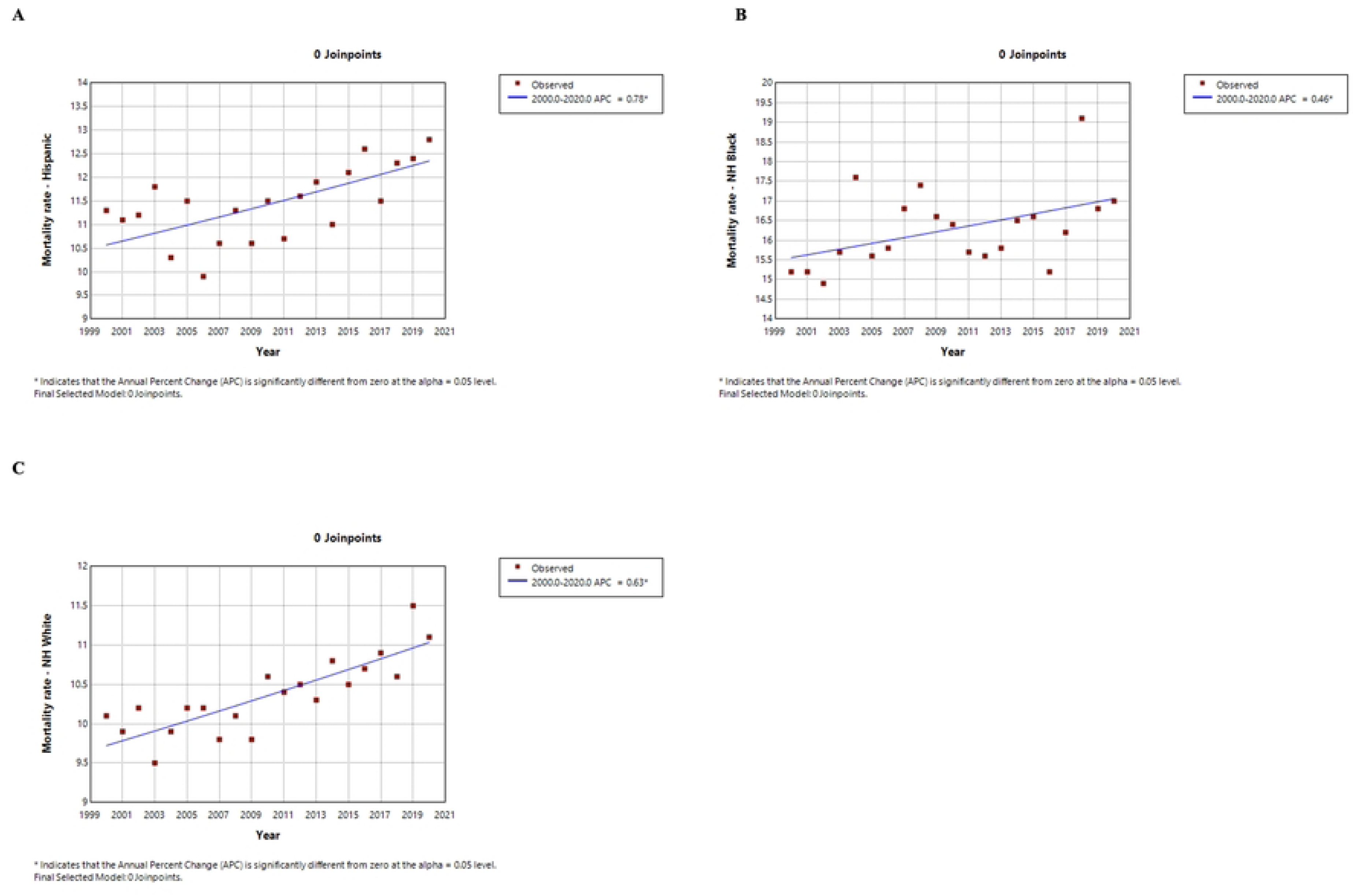
Joinpoint regression analysis of age-adjusted mortality rates of pancreatic cancer in Texas from 2000 -2020 by race/ethnicity. (A) Age-adjusted mortality rates for Hispanic (B) Age adjusted mortality rates for no-Hispanic Black. (C) Age-adjusted mortality rate for non-Hispanic White

Overall, Non-Hispanic Blacks had the highest age-adjusted mortality rates (13.8 per 100,000 persons) followed by Non-Hispanic White (10.4 per 100,000 persons) and then Hispanics (9.0 per 100,000).

## Discussion

We observed significant disparities in incidence and mortality trends across age, gender, and racial/ethnic groups in this population-based analysis of pancreatic cancer in Texas from 2000 to 2020. A total of 58,503 new cases of pancreatic cancer were reported with an average annual incidence rate of 12.2 per 100, 000 persons, with a slight male predominance and the highest incidence occurring in individuals aged 70–79 years, underscoring the disease’s strong association with advancing age [10]. These findings are consistent with prior studies showing that pancreatic cancer is primarily a disease of older adults, with median age at diagnosis in the U.S. typically above 70 years [11]. Males exhibited higher age-adjusted incidence rates than females (13.9 vs. 10.8 per 100,000), aligning with national trends reported by the Surveillance, Epidemiology, and End Results (SEER) Program [12]. Of particular note is the consistent disparity by race and ethnicity. Non-Hispanic Blacks had the highest incidence, followed by Non-Hispanic Whites and Hispanics, a pattern similarly reported in national epidemiologic studies [11, 12]. In addition, Tavakkoli et al [10] have also reported Non-Hispanic Blacks have the highest pancreatic cancer incidence and mortality rates predominantly in Midwest, Southeast and West South-Central states and across all age groups. Notably, Non-Hispanic Blacks have historically been shown to have both higher incidence and poorer survival, potentially due to a combination of biological factors, such as inherited mutations in genes like STK11, BRCA1, BRCA2, PALB2, CDKN2A and DNA repair genes [6], established risk factors such as cigarette smoking, diabetes mellitus, delayed diagnosis, and disparities in access to care [6, 13].

Temporal trends revealed a fluctuating but overall upward trajectory in pancreatic cancer incidence for both genders until around 2016, after which male rates slightly declined and female rates plateaued. Racial/ethnic trends showed sharper increases among Non-Hispanic Blacks, particularly between 2003–2004 and 2017–2019, while Hispanics exhibited more stable rates with a modest decline post-2017. These trends in incidence rates among non-Hispanic blacks could be partly explained by increased cigarette smoking and diabetes mellitus among black men, and obesity and heavy alcohol consumption among black women [13]. Edgar et al [14] also reported similar trends in pancreatic incidence among non-Hispanic blacks with the highest incidence rates during 2004 through 2008. Non-Hispanic Whites demonstrated a gradual increase until 2019, followed by a slight decrease. These variations may reflect underlying differences in environmental exposures, genetic predispositions, and modifiable risk factors such as smoking [15], obesity [16], and diabetes, which are more prevalent in certain subpopulations [17, 18]. Furthermore, socioeconomic barriers and health system inequalities likely contribute to the observed mortality patterns, where Non-Hispanic Whites accounted for the highest number of deaths, but Non-Hispanic Blacks had the highest incidence burden. These findings underscore the need for more equitable access to early detection strategies, public health interventions tailored to high-risk groups, and continued investigation into the underlying causes of these disparities.

Consistent with other studies [12], we observed that pancreatic cancer mortality rates differ substantially between male and female, and between blacks and non-Hispanic whites. Hispanics had the lowest mortality rate over the study period. Overall, males had a higher mortality rate, however we observed an increasing trend in mortality among females. Females demonstrated a consistent upward trend in mortality over the entire study period with an annual percentage change of 0.34% as compared to males where a statistically significant increase was observed from 2017 to 2020 with an annual percentage change of 2.76%. The acute rise in pancreatic cancer mortality in recent years could be attributed to the growing prevalence of its risk factors associated with globalization, urbanization, and economic development [19]. Huang et al [19] did a trend analysis on 48 countries for which he observed increasing trends in pancreatic cancer mortality among women in 14 countries for which the United States was included. This trend in women could be partly attributed to the higher prevalence of risk factors such as smoking, alcohol drinking, obesity, and metabolic syndrome [19, 20]. Additionally, rising incidence of pancreatic cystic neoplasm in females which is a new disease entity with increasing awareness in the past 2 decades may be attributed to higher prevalence of obesity and metabolic syndrome in females [19].

The higher mortality rates in blacks have been consistent with other studies.^10^ This may be due to a combination of factors such as poverty, insurance coverage, higher incidence rates, tumor biology, response to treatment and access to care [21]. However, we observed an increase in mortality among Hispanics, followed by non-Hispanic whites and Non-Hispanic blacks [APC: 0.78 vs 0.63 vs 0.46]. Hispanics experienced a rise in both incidence and mortality rates consistently over the study period. Gordon-Dseagu [22] found in their study that the incidence rates for the largest histologic type of pancreatic cancer, adenocarcinoma rose rapidly among Hispanic and Non-Hispanic white populations which may partially explain the rising mortality in these populations. Also, diabetes mellitus, a major risk factor for pancreatic cancer is rising in Hispanic and Non-Hispanic blacks compared to non-Hispanic whites [23, 24].

The persistent rise in both incidence and mortality, especially among minority groups, despite advancements in diagnostic imaging and increased awareness of risk factors, underscores the limitations of current detection and treatment strategies. Modifiable risk factors such as smoking, obesity, diabetes, and chronic pancreatitis remain more prevalent in some subpopulations and likely contribute to these observed disparities [17, 18, 25]. In addition, inability of diagnostic tools to detect early lesions, clinically silent early-stage pancreatic cancer, late-stage presentation and the lack of effective early detection tools continue to hinder meaningful improvements in survival [26]. However, there ae ongoing efforts being developed to explore possible ways for early detection. AI-enabled tools and machine learning in tandem with current imaging techniques are being developed [27, 28], blood tests are being developed as well for non-invasive early detection of pancreatic cancer [29].

There are some limitations in our study. First, Age-specific rates were suppressed when fewer than 16 cases were reported in specified categories, resulting in missing cases across some age groups. In addition, the use of administrative data may be subject to coding errors or incomplete information. The analysis did not account for important confounding factors such as health behaviors, comorbidities, and socioeconomic status, which can significantly influence pancreatic cancer incidence and outcomes. These factors may help explain the observed disparities and should be incorporated in future research to provide a more comprehensive understanding of pancreatic cancer patterns in Texas populations.

## Conclusion

Pancreatic cancer incidence and mortality rates are higher in males compared to females and non-Hispanic Blacks compared to Hispanic and non-Hispanic Whites. However, between 2000 - 2020, incidence and mortality rates have continued to rise particularly among females, Hispanics, and non-Hispanic blacks. Non-Hispanic whites have also seen an upward trend in incidence rates, particularly in 2009 -2017, and mortality rates during the study period. The higher incidence and mortality rates coupled with increasing trends in non-Hispanic Blacks could be partly explained by known risk factors for pancreatic cancer as well as significant inequities within the healthcare system. These disparities call for an integrated approach involving enhanced access to timely screening and treatment, improved patient education programs, and the removal of structural healthcare obstacles. Strategic focus on these fundamental components can help diminish existing outcome disparities and foster more equitable pancreatic cancer care across all racial and ethnic groups.

## Data Availability

The data underlying the results presented in the study are available from Texas Cancer Registry, https://www.cancer-rates.com/tx/

## Notes

### Competing Interest Statement

The authors have declared no competing interest.

### Funding Statement

The author(s) received no specific funding for this work.

### Author Declarations

Institutional Review Board (IRB) approval was not required for this study. This research utilized data from the Texas Cancer Registry, which is a publicly available, population-based cancer surveillance database containing only de-identified patient information. The Texas Cancer Registry data are considered exempt from human subjects research regulations under 45 CFR 46.104(d)(4) as the research involves the study of existing data that are publicly available and cannot be linked to specific individuals. No direct contact with patients or access to personally identifiable information occurred during this study. The research was conducted in accordance with the ethical principles outlined in the Declaration of Helsinki and followed all applicable guidelines for the use of population-based cancer registry data.

